# Effects of Age, Sex, Serostatus and Underlying Comorbidities on Humoral Response Post-SARS-CoV-2 Pfizer-BioNTech Vaccination – A Systematic Review

**DOI:** 10.1101/2021.10.10.21264825

**Authors:** Kin Israel Notarte, Abbygail Therese Ver, Jacqueline Veronica Velasco, Adriel Pastrana, Jesus Alfonso Catahay, Gian Luca Salvagno, Eric Peng Huat Yap, Luis Martinez-Sobrido, Jordi Torrelles, Giuseppe Lippi, Brandon Michael Henry

**Affiliations:** Faculty of Medicine and Surgery, University of Santo Tomas, Manila, Philippines; Service of Laboratory Medicine, Pederzoli Hospital, Peschiera del Garda, Italy; Section of Clinical Biochemistry, University of Verona, Verona, Italy; Lee Kong Chian School of Medicine, Nanyang Technological University, Singapore; Host-Pathogens Interactions and Population Health Programs, Texas Biomedical Research Institute, San Antonio, Texas, USA; Clinical Laboratory, Division of Nephrology and Hypertension, Cincinnati Children’s Hospital Medical Center, Ohio, USA

**Keywords:** COVID-19, mRNA vaccines, Pfizer-BioNTech, mRNA BNT162b2, immunoglobulins

## Abstract

With the advent of the Severe Acute Respiratory Syndrome Coronavirus 2 (SARS-CoV-2) pandemic, several vaccines have been developed to mitigate its spread and prevent adverse consequences of the Coronavirus Disease 2019 (COVID-19). The mRNA technology is an unprecedented vaccine, usually given in two doses to prevent SARS-CoV-2 infections. Despite effectiveness and safety, inter-individual immune response heterogeneity has been observed in recipients of mRNA-based vaccines. As a novel disease, the specific immune response mechanism responsible for warding off COVID-19 remains unclear at this point. However, significant evidence suggests that humoral response plays a crucial role in affording immunoprotection and preventing debilitating sequelae from COVID-19. As such this paper focused on the possible effects of age, sex, serostatus, and comorbidities on humoral response (*i*.*e*., total antibodies, IgG and/or IgA) of different populations post-mRNA-based Pfizer-BioNTech vaccination. A systematic search of literature was performed through PubMed, Cochrane CENTRAL, and Google Scholar. Studies were included if they reported humoral response to COVID-19 mRNA vaccines. A total of 32 studies was identified and reviewed, and the percent difference of means of reported antibody levels were calculated for comparison. Findings revealed that older individuals, the male sex, seronegativity, and those with more comorbidities mounted less humoral immune response. Given these findings, several recommendations were proposed regarding the current vaccination practices. These include giving additional doses of vaccination for immunocompromised and elderly populations. Another recommendation is conducting clinical trials in giving a combined scheme of mRNA vaccines, protein vaccines, and vector-based vaccines.

## INTRODUCTION

Over 200 million infected cases and over 4.6 million Coronavirus Disease 2019 (COVID-19) deaths have been reported globally [1]. The rapid transmissibility of Severe Acute Respiratory Syndrome Coronavirus 2 (SARS-CoV-2) including variants being monitored (VBM) and variants of concern (VoC), have sparked fear worldwide and forced many countries to deal with repeated surges in confirmed cases and deaths [2]. To control the pandemic, innovative therapeutic strategies have been formulated, with the specific aim to avert clinical outcomes and limit morbidity, disability, and death associated with COVID-19 [3]. A SARS-CoV-2 vaccination program that is cost-effective, safe, and efficacious is also implemented globally. Among the different types of COVID-19 vaccines, the utilization of a new generation of mRNA-based vaccines is unprecedented, and showed high efficacy to trigger immunoprotective humoral response [4]. Despite their effectiveness in reducing the risk of infection and clinical deterioration, the considerable inter-individual heterogeneity in post-vaccine immune response has been increasingly observed in specific populations, particularly among the elderly and immunocompromised individuals [5,6]. To account for low vaccine responders or individuals with less effective production of neutralizing antibodies, this systematic review is focused on determining the possible effects of age, sex, serostatus, and underlying comorbidities on humoral response of recipients of mRNA-based vaccines.

## METHODS

### Search Strategy and Eligibility Criteria

A systematic literature search was conducted to identify studies reporting the factors affecting humoral response of individuals who received the mRNA vaccines. As shown in Figure 1, a comprehensive search was carried out in PubMed, Cochrane CENTRAL, and Google Scholar for articles published from January to end of July 2021. The search keywords include “age”, “sex”, “seropositivity”, “comorbidities”, “humoral response”, “mRNA vaccine”, “Pfizer-BioNTech”, and “mRNA BNT162b2” which resulted in 150 journal articles. For the inclusion criteria, articles reporting the following data were considered: (1) total IgG or IgA or neutralizing antibody titers, (2) quantitative antibody tests, (3) mRNA-based vaccines, (4) articles reporting time points for data extraction, (5) articles available in the English language, and (6) randomized controlled, cohort, preprint or published papers as long as they provide extractable data given the limited papers available for this novel disease and the mRNA vaccine. Exclusion criteria were IgM response and cell-mediated immunity, the former because it is now increasingly clear that IgM plays a minor role against COVID-19 (it does not always appear, wanes very early and has lower neutralizing potential), the latter because we only focused on humoral immunity.

**Figure 1.**
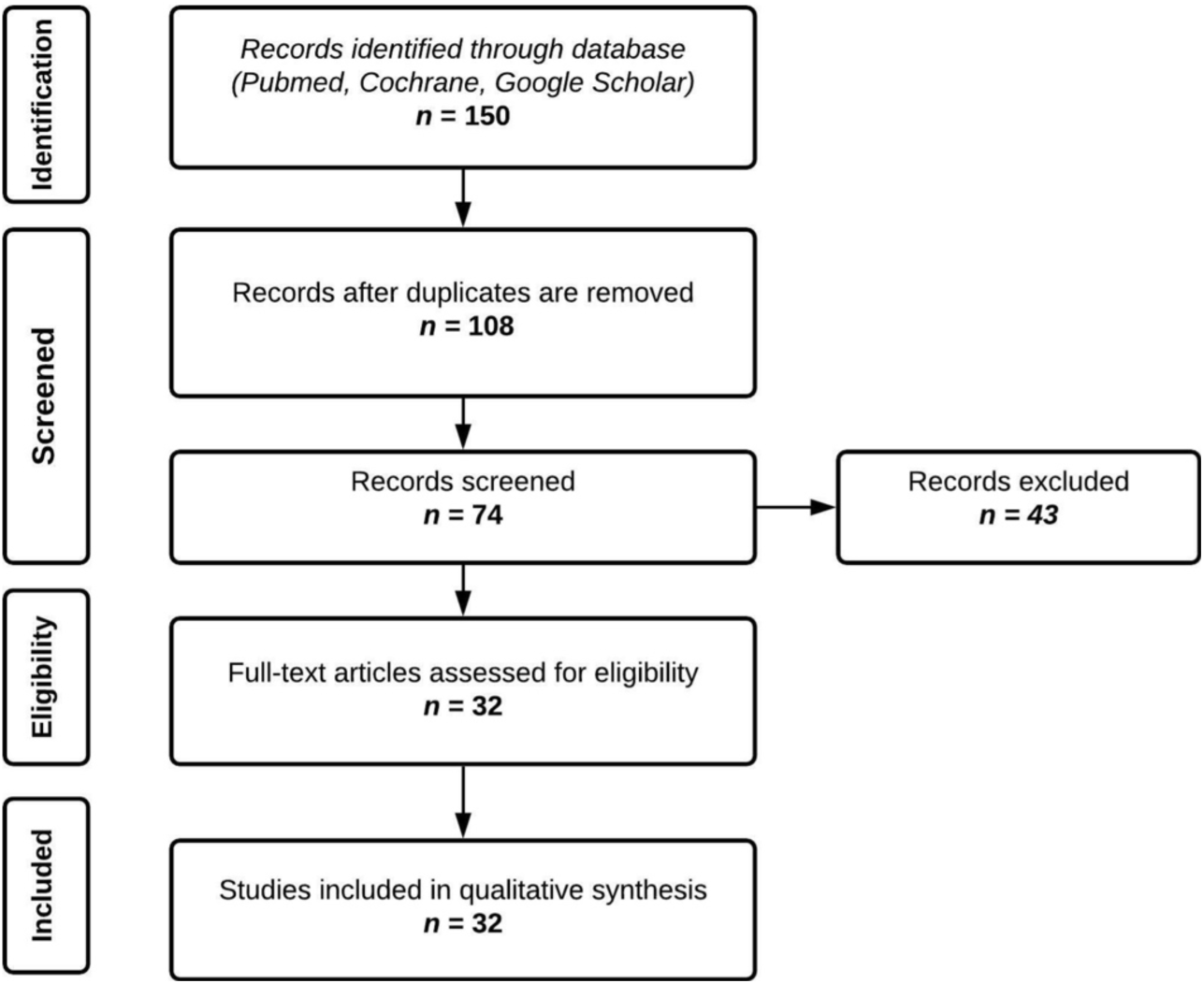
Screening and appraisal of journal articles for inclusion in the systematic review.

The search results were tabulated, and duplicates were removed. Full text of each article was retrieved and assessed for final eligibility independently by any two reviewers. Any disagreement was resolved by consensus of all authors.

### Data Extraction

Descriptive and outcome data were extracted from the included studies. The extracted data includes the type of vaccine, country of origin, sample size, age range or median age of the population, type of immunoglobulin measured, and percent mean difference between the control and factors affecting humoral immune response observed in the various studies. Additional data was requested from the original study authors when necessary.

### Data Analysis

Due to significant heterogeneity in the assays used to probe antibody titers, we standardized antibody measurements reported in the articles that were included using a percent difference of means computed using the formula:

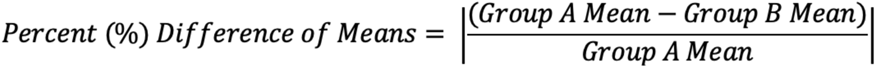

This enables the comparison of laboratory results regardless of the units of measurements and the diagnostic tools for quantification of antibody titers.

### Scope and Limitations

The demographic parameters used in this study were limited to age, sex, serostatus, and comorbidities such as hemodialysis or end-stage renal disease (ESRD), transplant recipients, and metabolic derangements including obesity, hypertension, and smoking. These factors were analyzed independently of each other, with the exception of concurrent effects of age with comorbidities.

In addition, only Pfizer-BioNTech (mRNA BNT162b2) vaccine was used in this study as it was the leading vaccine utilized worldwide and due to the wide range of available studies at the time of writing. Moreover, the authors utilized both preprint and published journal articles written in the year 2021, with at least a record of 10 patients and above, used mRNA vaccines and quantitative antibody tests, and reported IgG or IgA to determine humoral response. No data on cell mediated immunity was included and all time points shown in the data were in reference to the first dose after vaccine administration.

## RESULTS

A total of 32 articles were included in our analysis. These were divided into four categories, humoral response influenced by (1) age, (2) sex, (3) baseline serostatus (*i*.*e* seropositive or seronegative), and (4) presence of comorbidities. Seven articles were included under the age category, three under the sex category, 12 under serostatus, and 18 under the comorbidities category. The comorbidity category was further subdivided into four classes: hemodialysis or end stage renal disease (5 articles), cancer and autoimmune diseases (6 articles), transplant patients (4 articles), and metabolic derangements (3 articles). The articles reviewed under each category were non-exclusive, as most studies analyzed their samples with at least two of the mentioned factors.

Tables 1-4 provide a summary of the following trends observed in this study. In general, mRNA vaccines were able to mount efficient antibody responses; however, the level of titers produced varied according to the factors of age, sex, serostatus, and comorbidities. The rate at which antibodies produced decline over time is also influenced by the aforementioned factors. Older individuals, the male sex, seronegativity, and those with more underlying comorbidities mounted less humoral immune response. Aging, in particular, is a significant aggravating factor in the decline of humoral response among recipients with underlying comorbidities, especially when compounded with immunosuppressive medications.

**Table 1.**
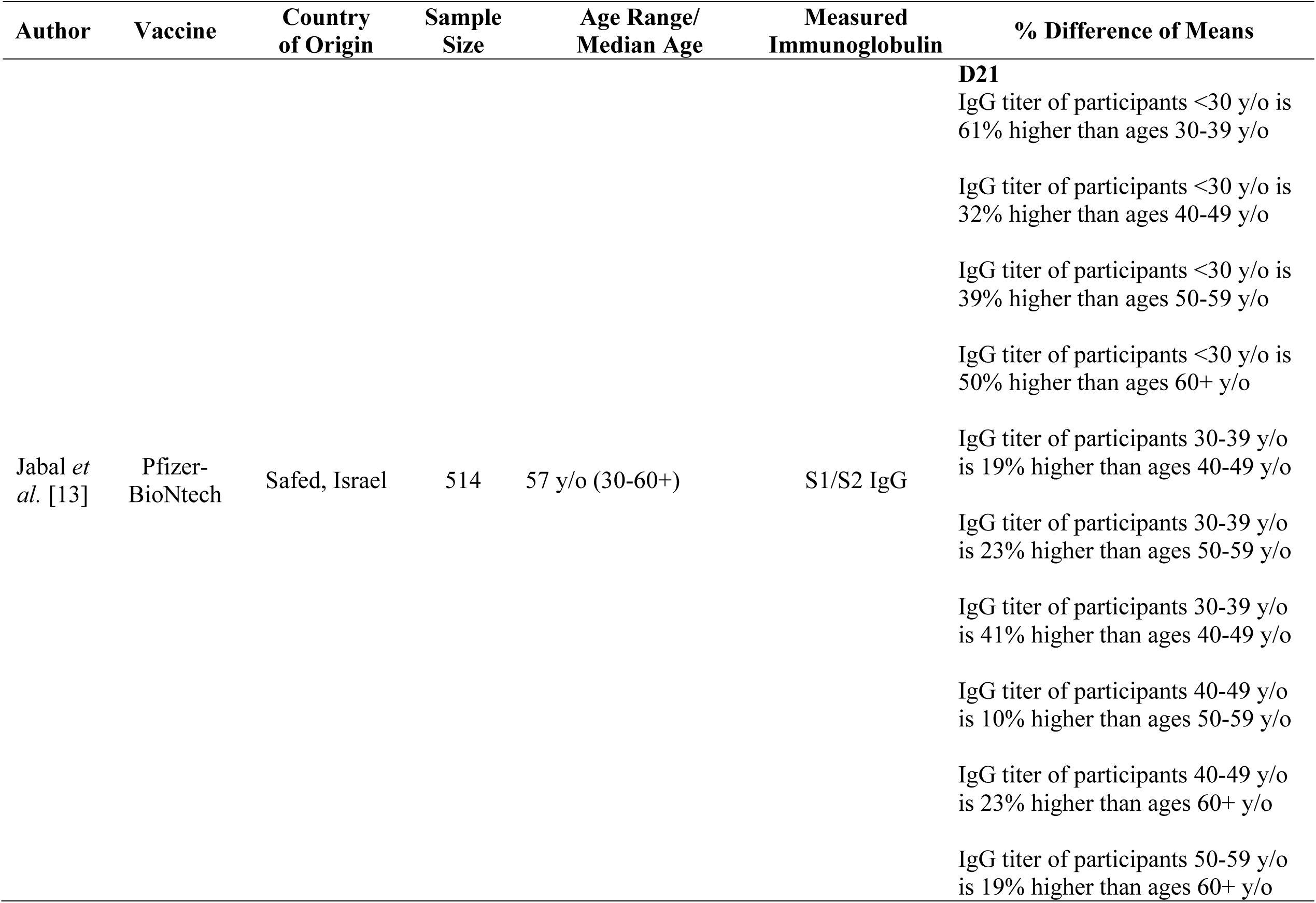
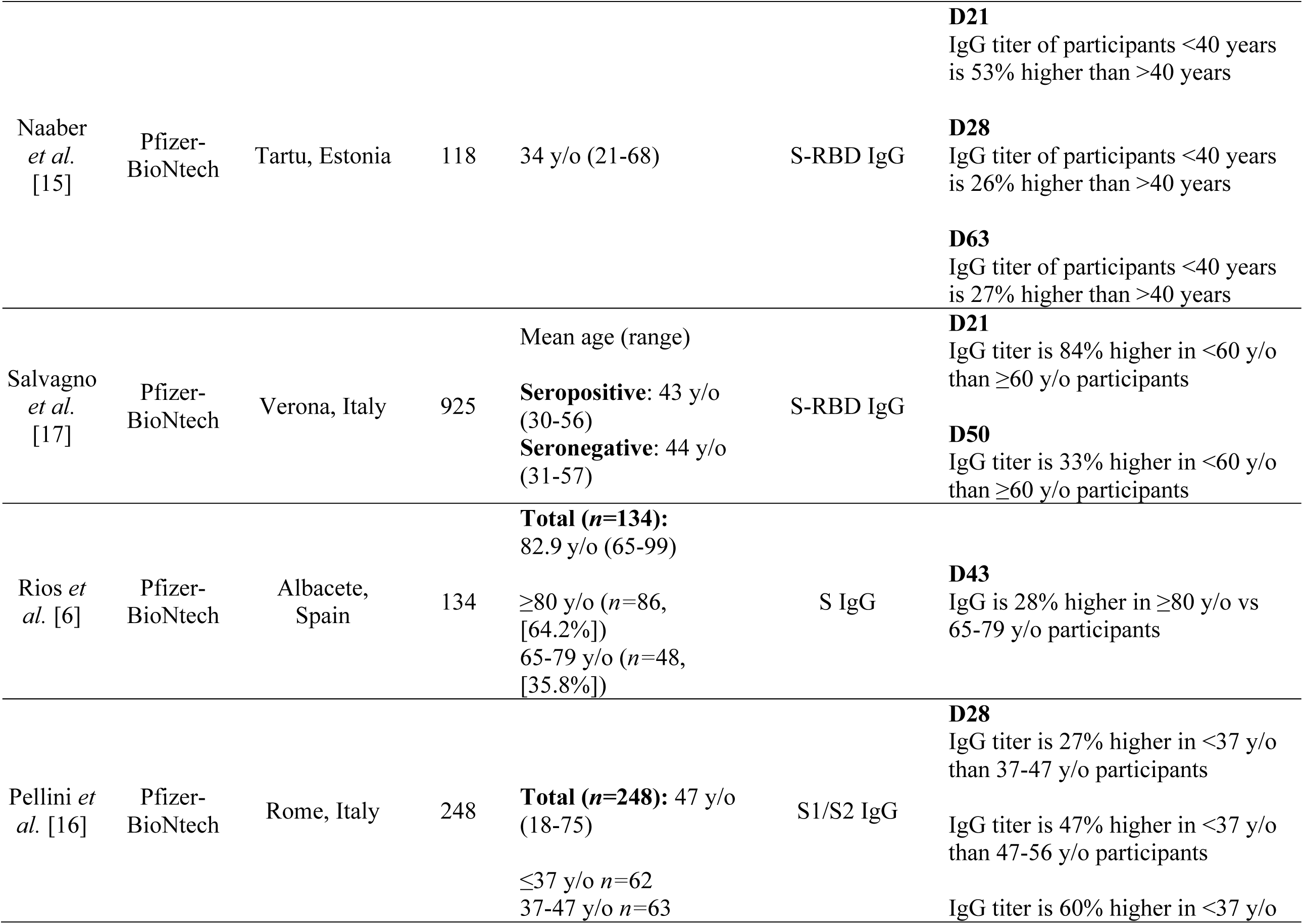
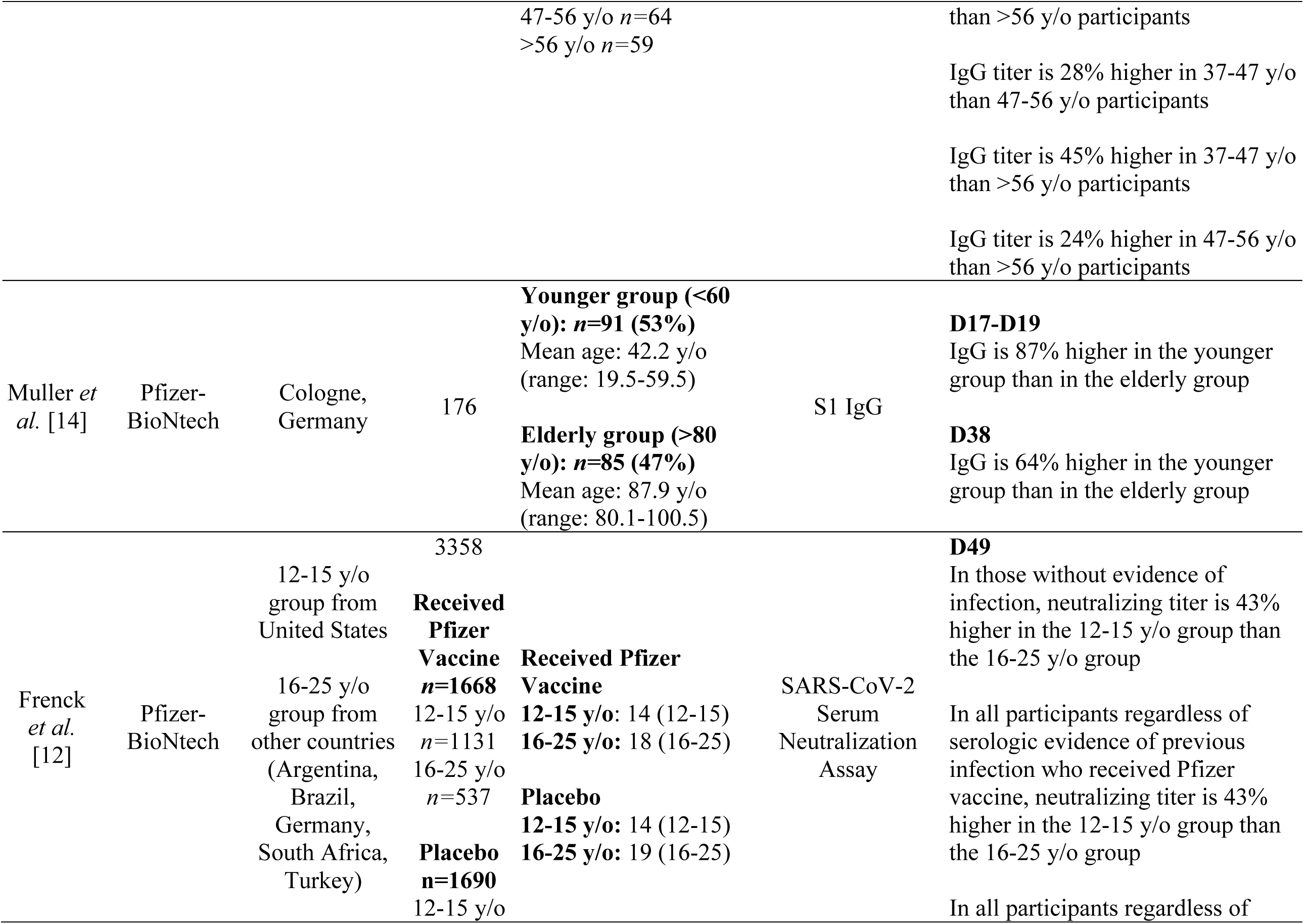
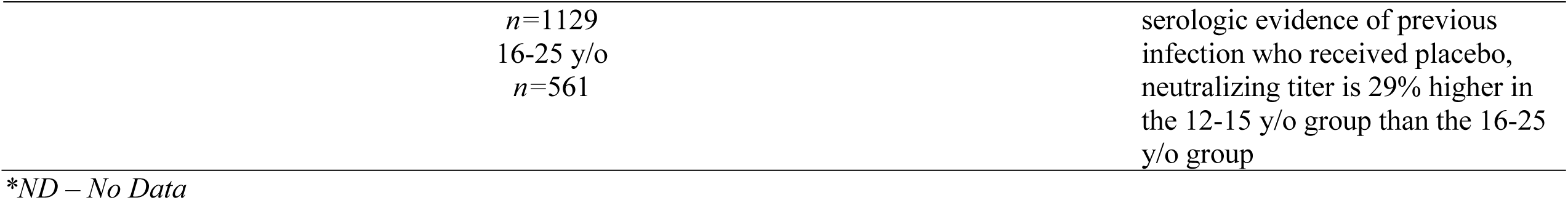
Effect of age in humoral response following Pfizer-BioNTech (mRNA BNT162b2) vaccine administration.

## DISCUSSION

### Factors Affecting Humoral Response

#### Age

A critical factor that makes the elderly more susceptible to infectious diseases is immunosenescence or the decline of immune system functionality as people age [7]. Immunosenescence has been linked with diminished response to vaccination and could therefore influence the success of vaccination [8]. There is clear evidence that the decline in adaptive immunity results in dramatically reduced vaccine responses and vaccine longevity in older adults [7]. This is well-documented with influenza A/H1N1 vaccination, where age negatively correlates with humoral immunity [9,10]. Like any other vaccine, there is accumulating evidence that immunosenescence could also impact the effectiveness of COVID-19 mRNA vaccines, and may hence be less protective to the elderly [11].

Several journal articles reviewed in Table 1, regardless of age stratification, reported that younger individuals developed a greater antibody response after vaccination compared to older individuals [12–17]. Meanwhile, Rios *et al*. reported no association between age and antibody response [5]; however, this article may be limited by the characteristics of its sample size. The study used residents of long-term care facilities, with variable disability and frailty profiles, who had a mean age of 82.9 years, with a range of 65-99 years.

Nevertheless, the findings of most of the studies were consistent with the current knowledge that there is diminished humoral response among the elderly (>65 years), owing to qualitative differences in memory B cells and plasma cells, and expansion of a pro-inflammatory subset of B cells [16]. Elderly individuals were noted to have decreased vaccine-specific antibody titers, and thus were more likely to be non- or low responders [15]. In addition, elderly individuals had more rapidly waning antibody levels [15]. This difference in antibody response was most prominent following the first dose of COVID-19 mRNA, but subsequently decreased over time, especially after the second dose [14,15,17].

Interestingly, the relationship between age and IgG or IgA antibody response was not limited to the elderly. Individuals across all age groups demonstrated this trend. Young individuals (12-64 years) consistently produced increased antibody titers compared to their older counterparts. This difference was even more pronounced between groups with large age gaps, further emphasizing the effect of age on antibody response.

These differences in antibody response have practical implications in COVID-19 vaccination programs. It highlights the importance of a second (or even a third) dose in order to boost the protective response in older individuals [15]. It also highlights the need to individualize vaccination programs and create strategies to account for possible age-related limitations for COVID-19 vaccination [14].

#### Sex

Females develop a greater antibody response due to hormonal differences compared to males, which regulates both adaptive and innate immune responses, with estradiol and testosterone having enhancing and suppressive effects, respectively [19]. However, levels of sex hormones change with age. Thus, after menopause the drop in estradiol levels enhances immunosenescence [19]. Studies in childhood vaccination enables research that focus on sex-dependent response aside from sex hormones that increase after puberty. Such is the case in studies on Measles, Mumps, and Rubella (MMR) and Diphtheria, Pertussis, and Tetanus (DPT) vaccines [20]. These suggest that genetic factors may play a role. The X chromosome expresses more genes, many of which influence immunity. This includes microRNAs (miRNAs) that are known to modulate immunity [20]. Several studies suggest that a similar trend is also observed among COVID-19 mRNA vaccines, noting a higher humoral response and adverse events among women.

Among the articles reviewed in Table 2, two studies showed a direct relationship between sex and humoral response. Both studies by Jabal *et al*. and Pellini *et al*. reported that the female sex is generally superior to male in terms of production of IgG in Day 21 and Day 28 post-vaccination of Pfizer-BioNtech, respectively [13,16]. These data further strengthen the previous studies of Ciarambino *et al*. that female gender has generally decreased susceptibility to viral infections from protection given by X chromosome and/or sex hormones [21]. The X chromosome provides females greater inflammatory, antiviral, and humoral immune responses compared to males. In addition, estrogen, a key hormone in females, plays a significant part in immune regulation.

**Table 2.** Effect of sex in humoral response following Pfizer-BioNTech (mRNA BNT162b2) vaccine administration.

| Author | Vaccine | Country of Origin | Sample Size | Age Range/ Median Age | Measured Immunoglobulin | % Difference of Means |
| --- | --- | --- | --- | --- | --- | --- |
| Salvagno *et al.* [17] | Pfizer-BioNtech | Verona, Italy | 925 | **Seropositive:** Range, 43 +/- 13 years<br>**Seronegative:** Range, 44 +/- 13 years | S-RBD IgG | **D30**<br>Among all participants, IgG titer of female participants is 13% higher than males<br>Among baseline seronegative participants, IgG titer of female participants is 16% higher than males |
| Pellini *et al.* [16] | Pfizer-BioNtech | Rome, Italy | 248 | 47 y/o (range 23-69 y/o) | S1/S2 IgG | **D28**<br>IgG titer of female participants is 37% higher than males |
| Jabal *et al.* [13] | Pfizer-BioNtech | Safed, Israel | 514 | 57 y/o (range 30 - 60+) | anti-N IgG | **D21**<br>IgG titer of female participants is 15% higher than males |

\*ND – No Data

Furthermore, studies by Ma *et al*. showed that estrogen can directly inhibit SARS-CoV-2 replication by regulating cell metabolism and maintaining cell integrity through genetic modification and improving metabolic function, reducing incidence of SARS-CoV-2 infection [22]. In contrast, testosterone suppresses immune functions by acting on androgen receptors and immune cell activity thereby decreasing inflammation and promoting anti-inflammatory responses. Thus, females have baseline physiologic advantage in mounting immune response than males.

### Serostatus

Vaccines act by triggering the body’s immune response, leading to development of humoral and cellular immune responses [17]. Several studies provided evidence that individuals previously infected with SARS-CoV-2 develop early antibody responses right after the primary infection, resulting in inter-individual heterogeneity in post-vaccine immune response [23,24]. The preservation of B-cell mediated memory immunity from patient’s previous SARS-CoV-2 infection has been theorized to be the primary cause of the boost-like immune response after COVID-19 vaccination in seropositive individuals [13,25].

Publications in Table 3 showed a robust accelerated, humoral immune response after the first vaccine dose. However, results of antibody titers after the second dose displayed different trends. In two separate studies, antibody titers of seropositive individuals are multiple folds higher than those baseline seronegative subjects after the second vaccine dose [17,26]. On the other hand, some publications stated that there is no significant difference between the two groups after the second dose [27,28,29]. As a result, different recommendations can be gathered from different sources. Kelsen *et al*. recommended that subjects with prior COVID-19 may require only a single dose of vaccine [29], and eventually a second dose only when the antibody levels decline significantly (*i*.*e*., typically, after 12 months), or in case new VBM and VoC, characterized by the so-called “escape mutations”, become endemic. This is in contrast with the recommendation of Demonbreun *et al*. stating that one vaccine is not enough to produce strong protection against SARS-CoV-2 infection among most people previously infected [30]. Demonbreun *et al*. reported that the humoral response of the seropositive group was significantly lower than the response of the PCR (+) group. The study defined the seropositive group based on the presence of anti-RBD IgG antibodies, while participants who tested positive for SARS-CoV-2 on a clinical molecular diagnostic test for acute infection any time prior to vaccination were categorized as recovered COVID-19 under the PCR (+) group [30]. Yan *et al*. elaborated that the difference among anti-SARS-CoV-2 titers may be attributed to the higher initial amounts of viral antigens in cases of severe COVID-19. It may also be the result of an excessive immune response, selective B cell plasmablast amplification, and enhanced and prolonged B cell receptor stimulation in patients with severe COVID-19 [31].

**Table 3.**
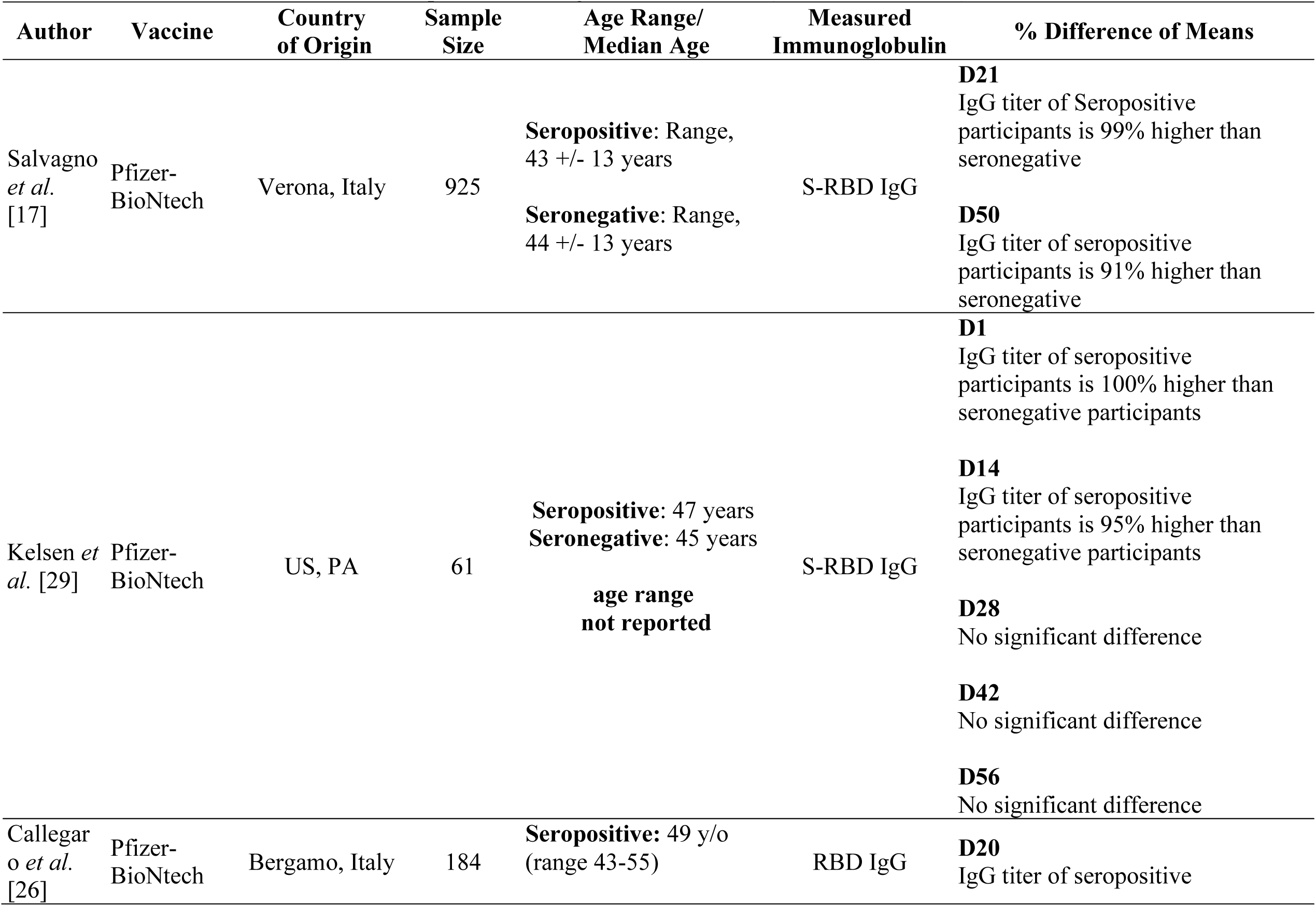
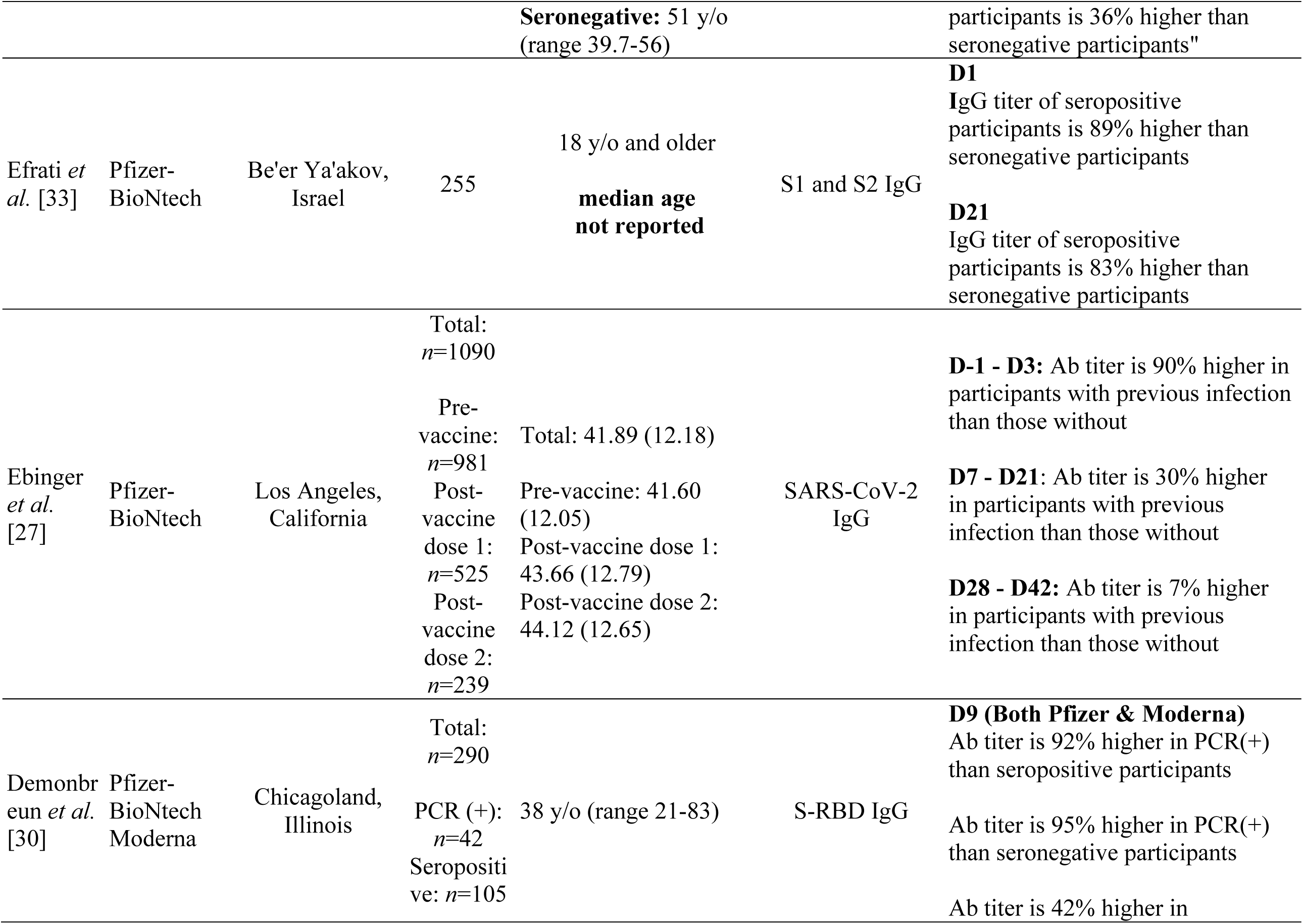
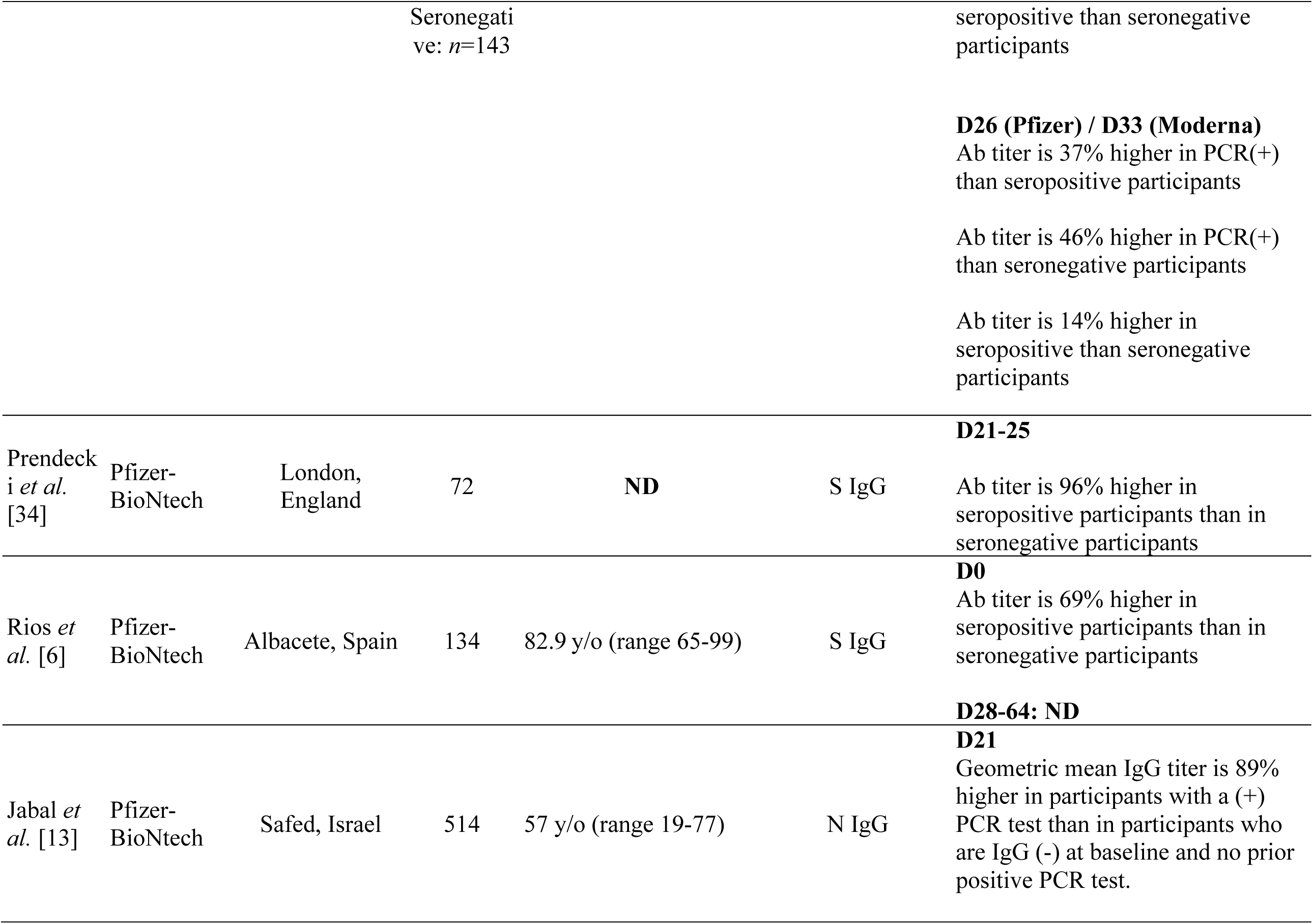
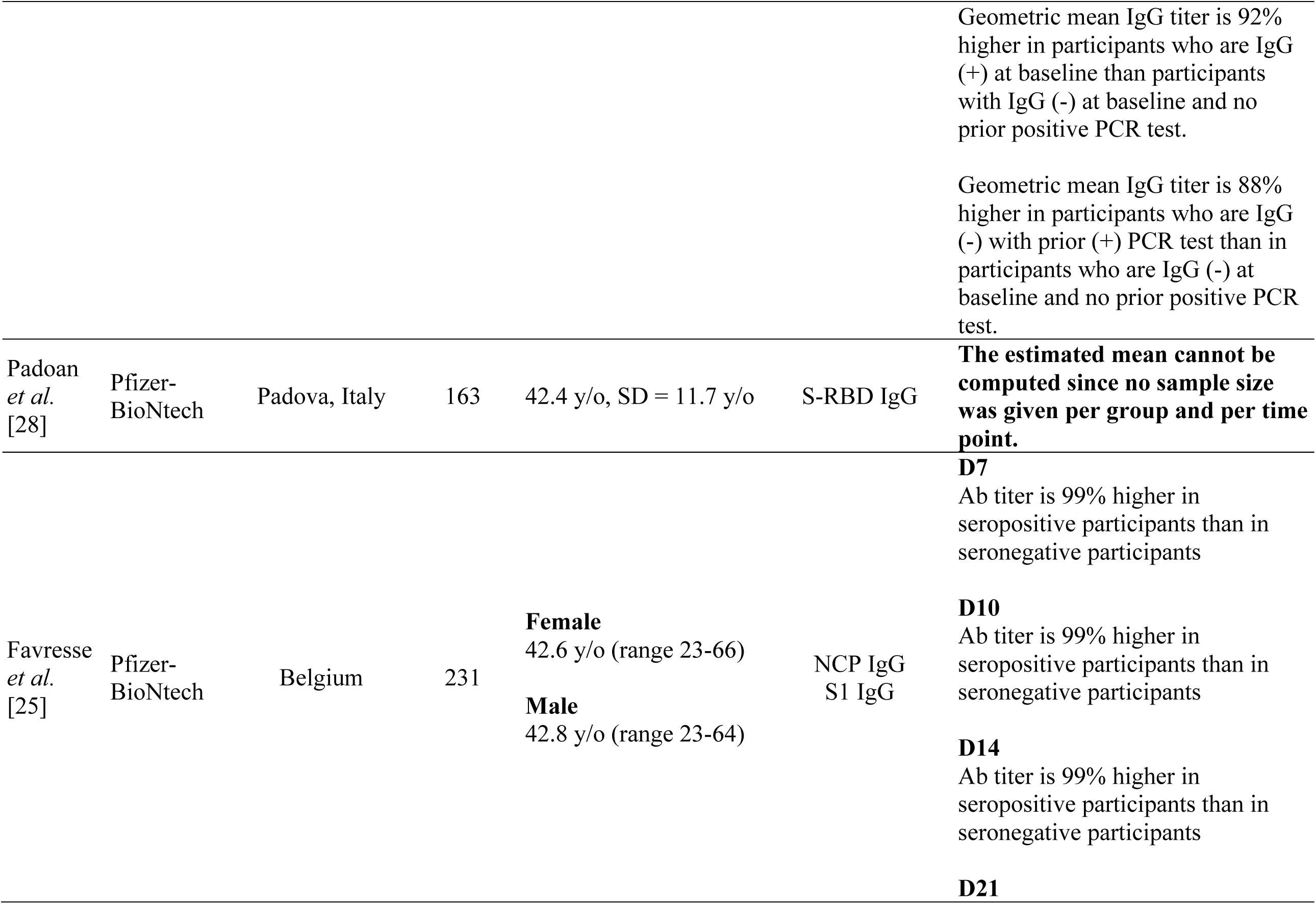
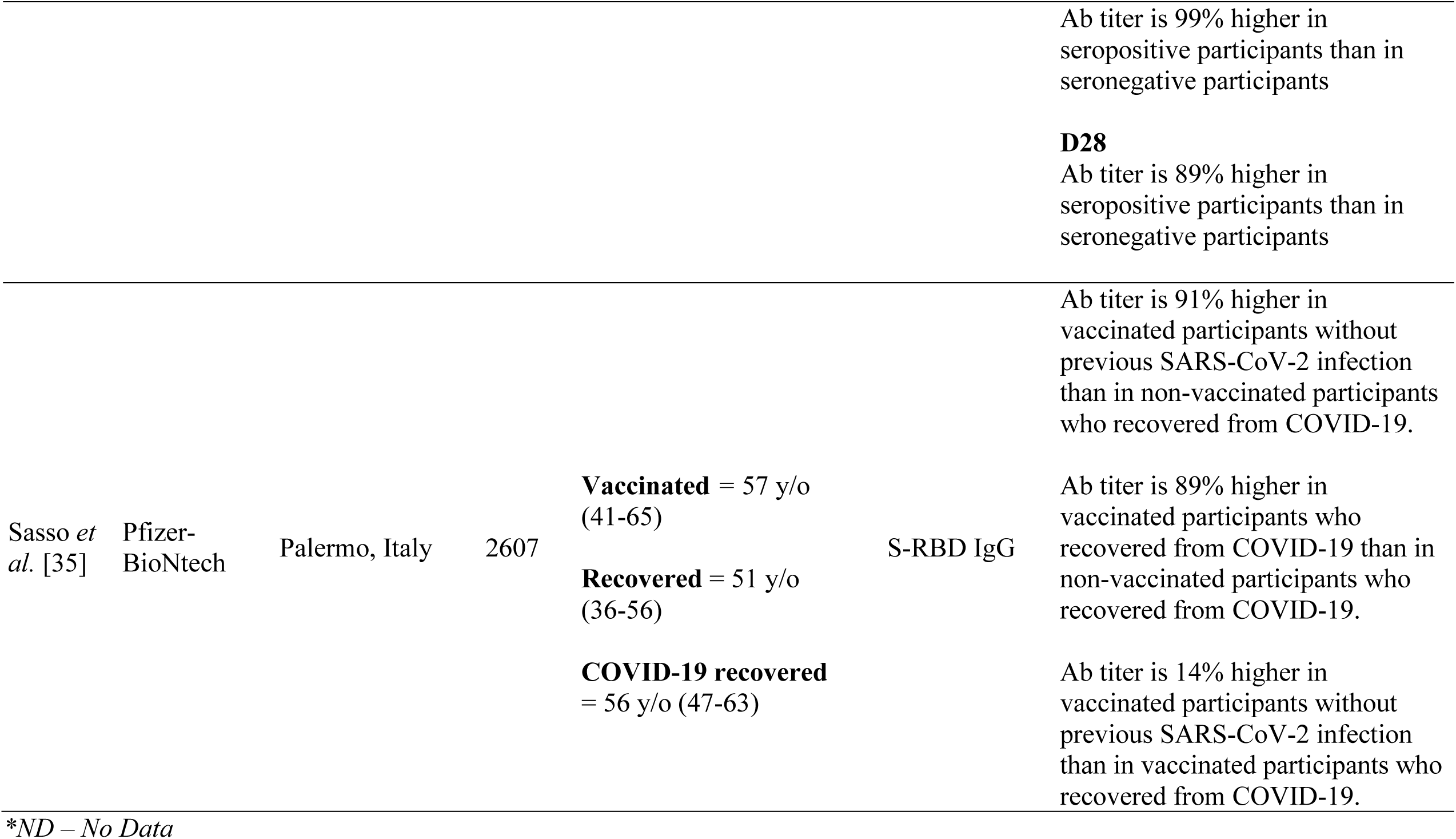
Effect of serostatus in humoral response following Pfizer-BioNTech (mRNA BNT162b2) vaccine administration.

Therefore, proper investigation is necessary so that the heightened immune response among seropositive individuals may be used to incite strategic change among vaccine distribution, promoting faster vaccine dispensing and better vaccine allocation, especially to high-risk populations [32].

### Comorbidities

The presence of comorbidities, such as chronic kidney diseases (CKD), diabetes mellitus (DM) and cardiovascular disease (CVD), are clinical risk factors significantly associated with poorer prognosis in patients with COVID-19 [34]. This is greatly attributed to the disease mechanisms resulting in metabolic disorders that impair lymphocyte and macrophage functions, hence also negatively affecting the immune response after COVID-19 vaccination [37,38]. To highlight the effect of comorbidities in humoral response of individuals receiving the COVID-19 mRNA vaccine, Rios *et al*. reported that immune response in the elderly with higher number of comorbidities is blunted by 30% as compared to those with lesser comorbidities [6]. Thus, the high prevalence of people with comorbidities, combined with increasing age (>60 years) as seen in Table 4, shows that the presence of both factors may aggravate the already blunted response in affected individuals, as aging accelerates the rapid decline in humoral immunity. [39]. The following sections outline the different comorbidities that may affect humoral response following COVID-19 mRNA vaccination, as highlighted in Table 4.

**Table 4.**
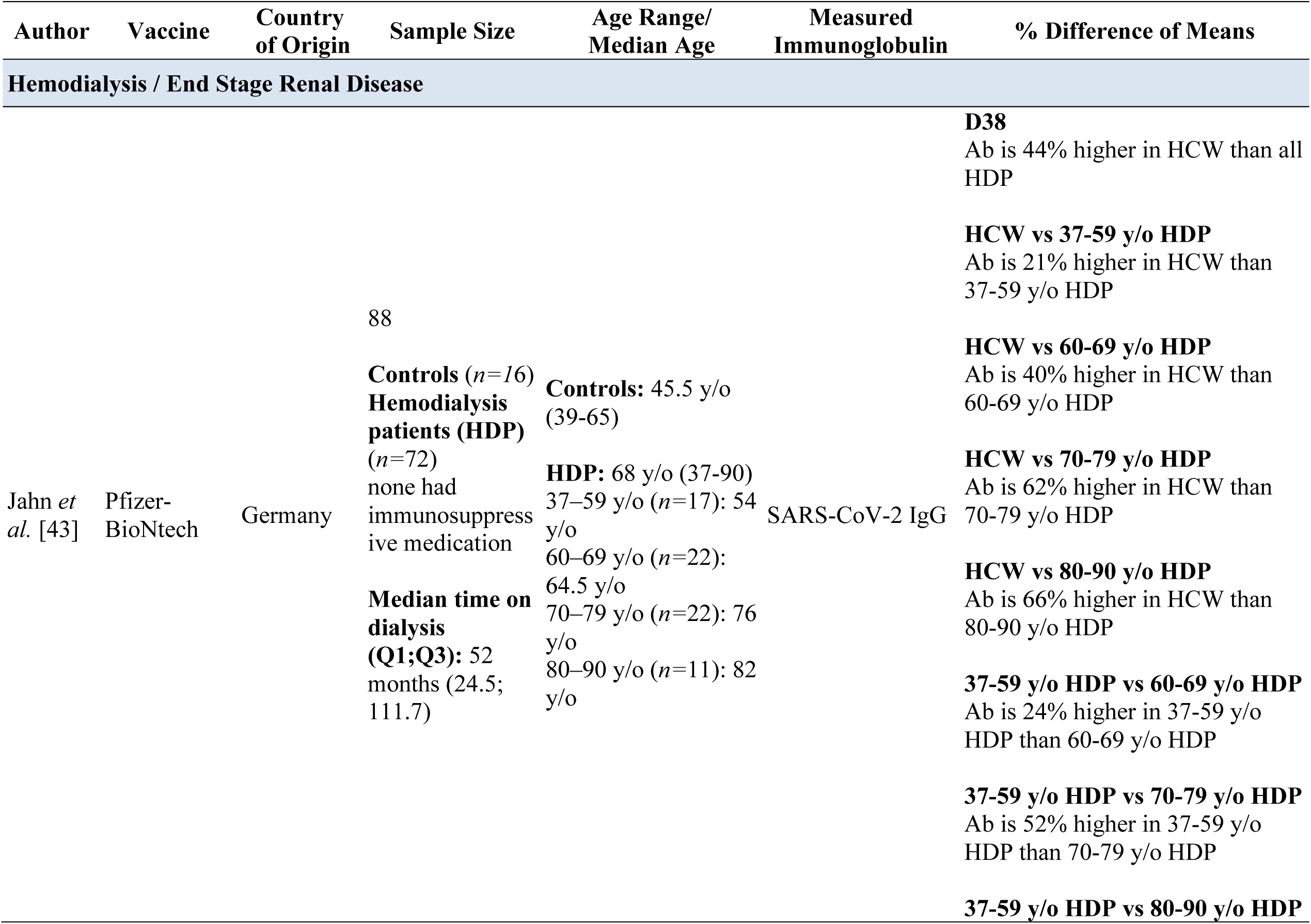
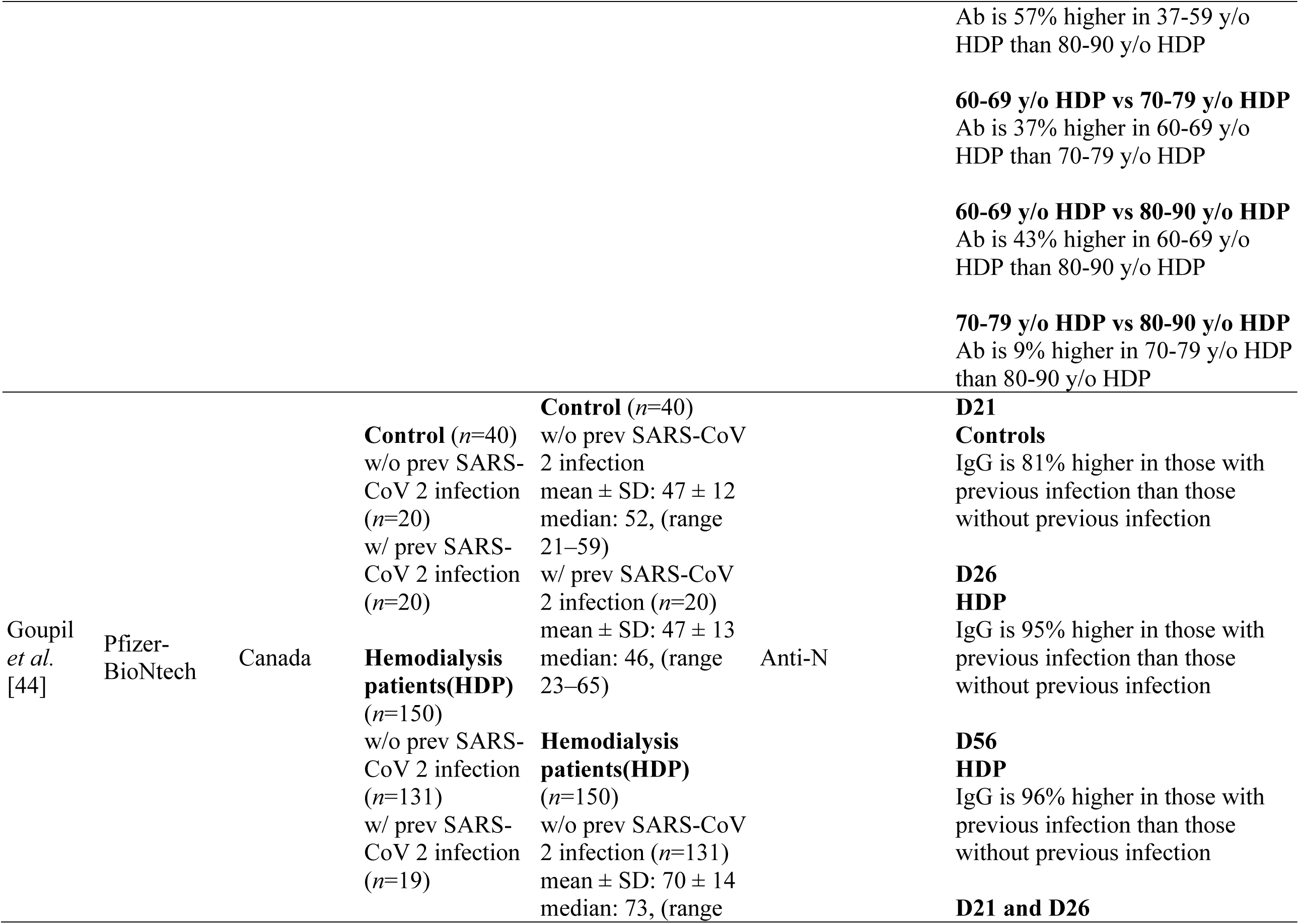
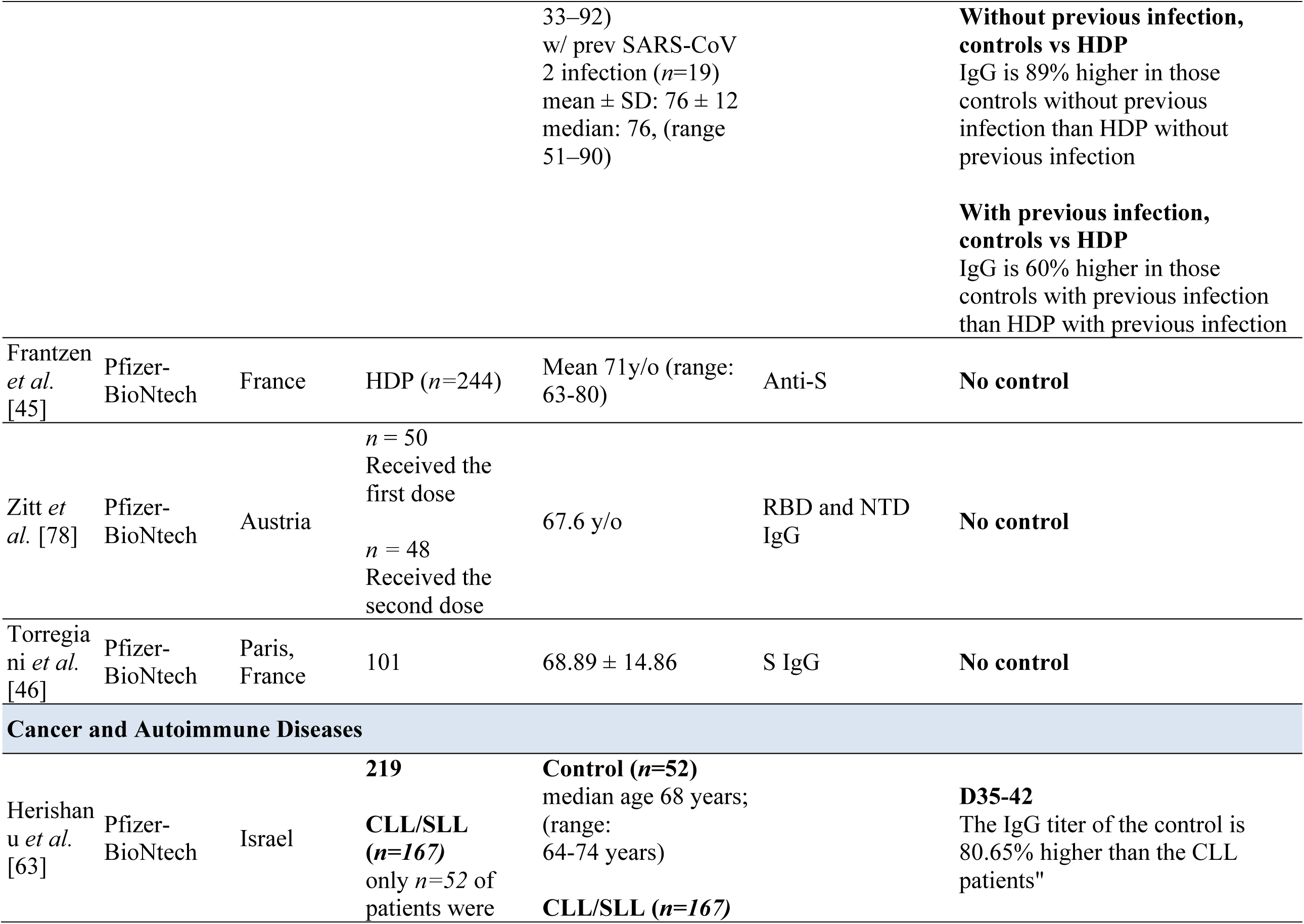
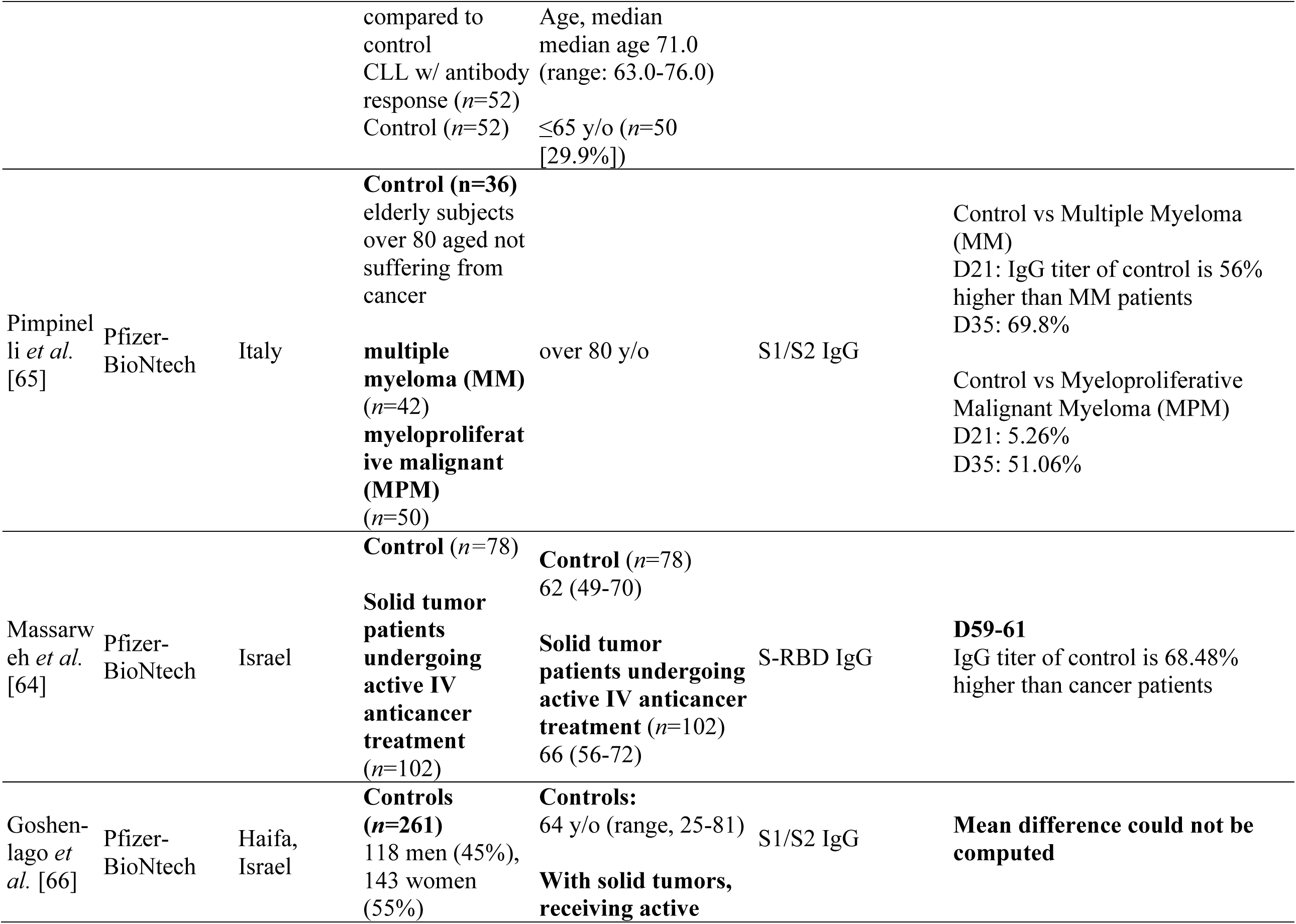
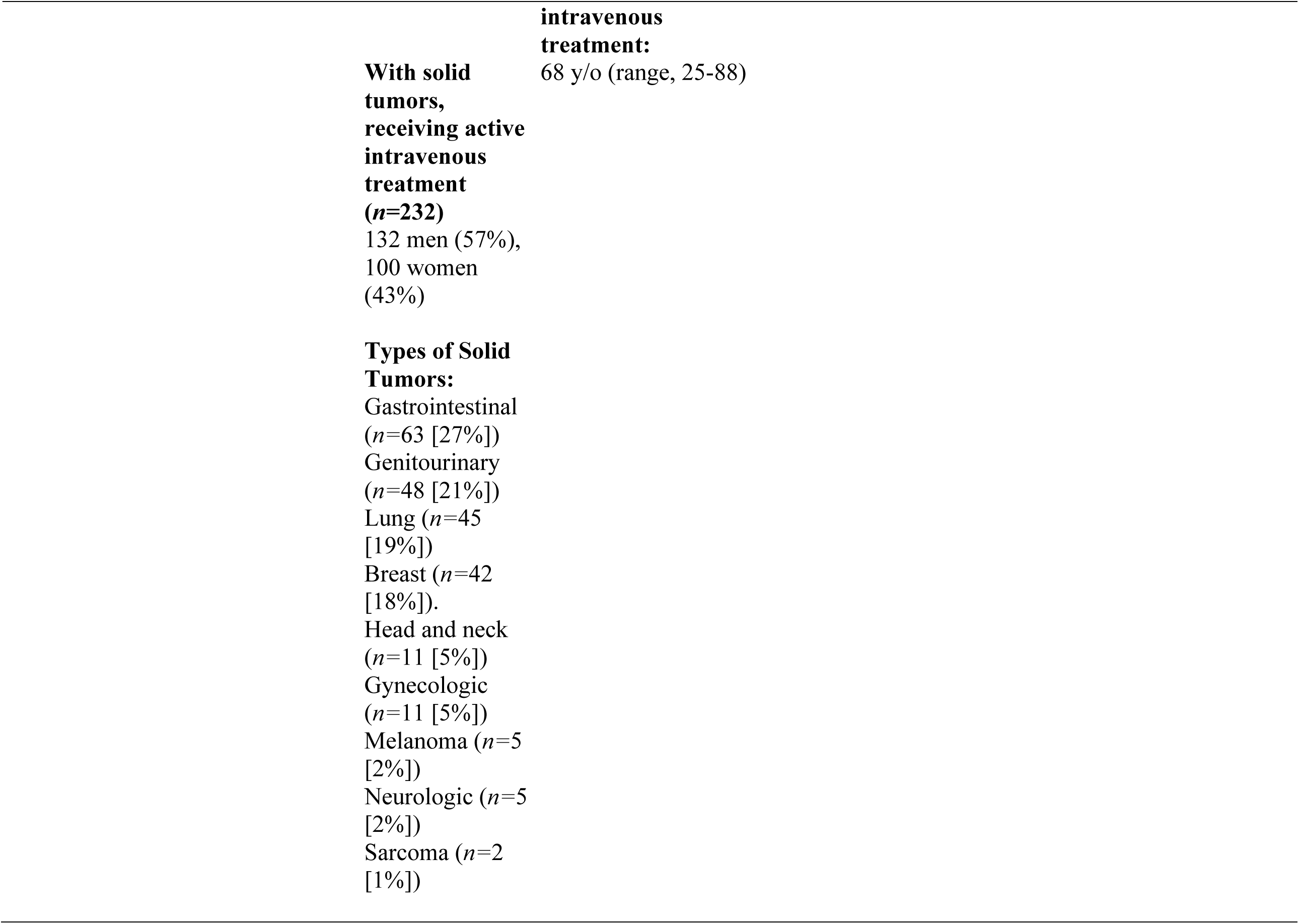
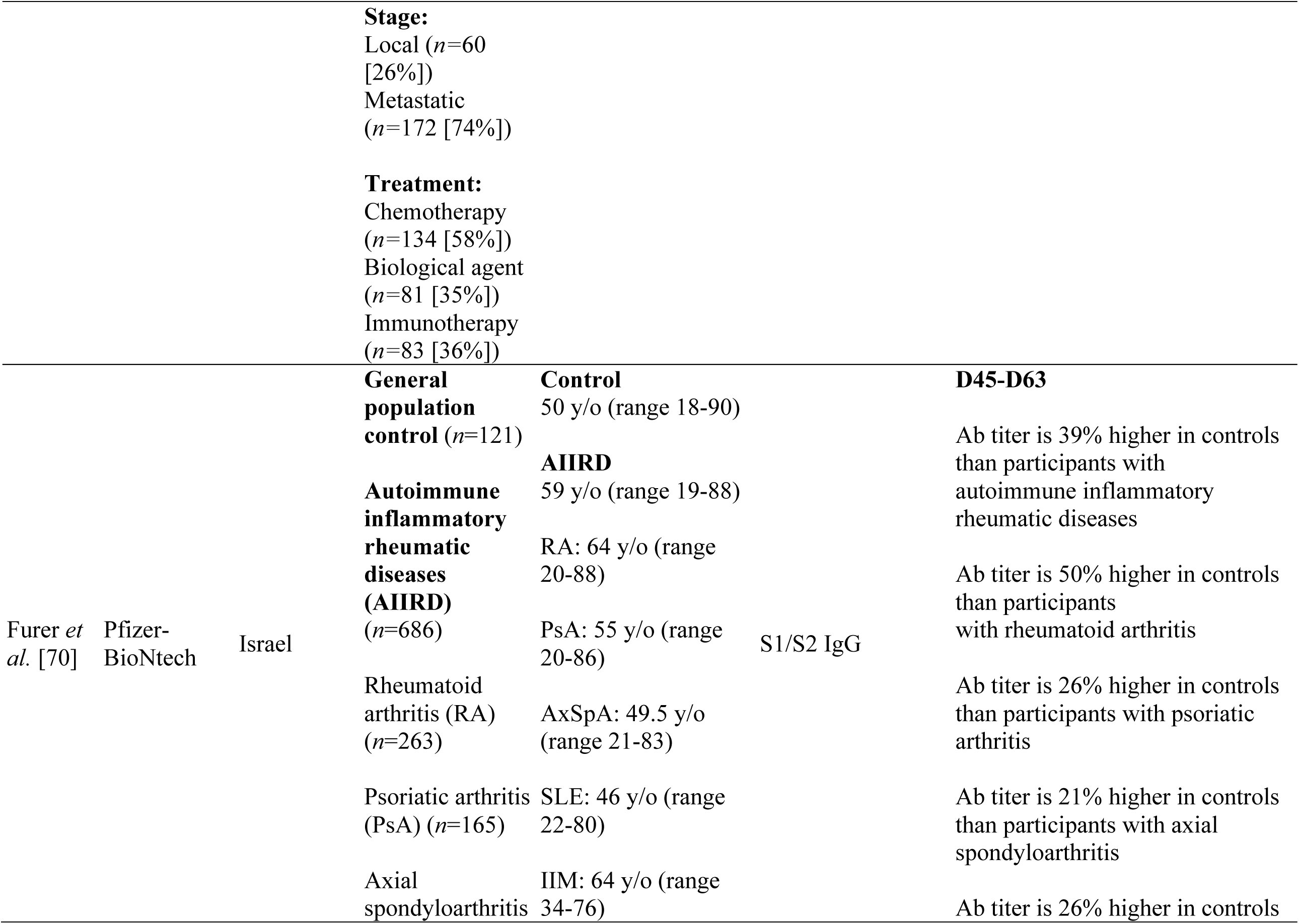
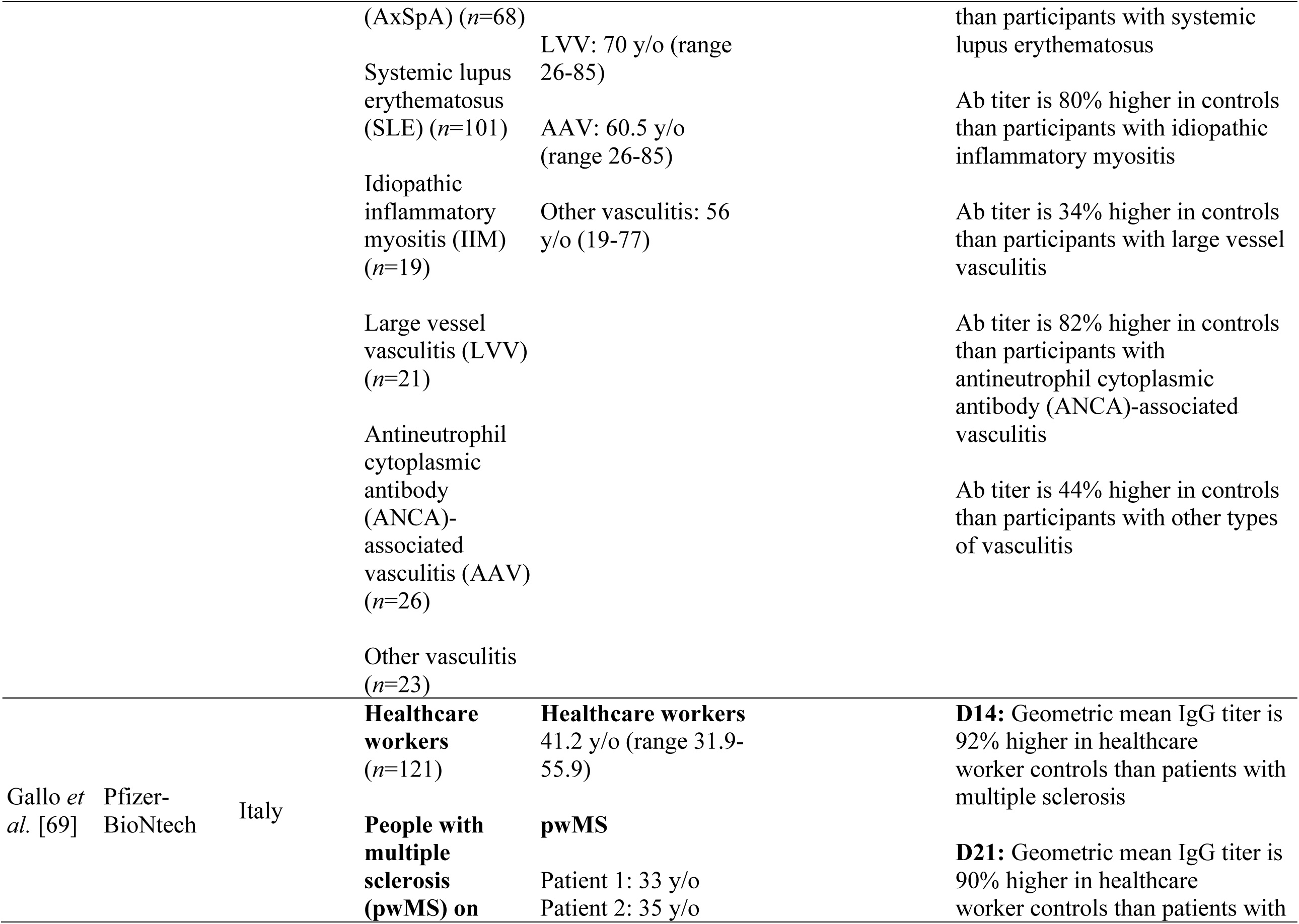
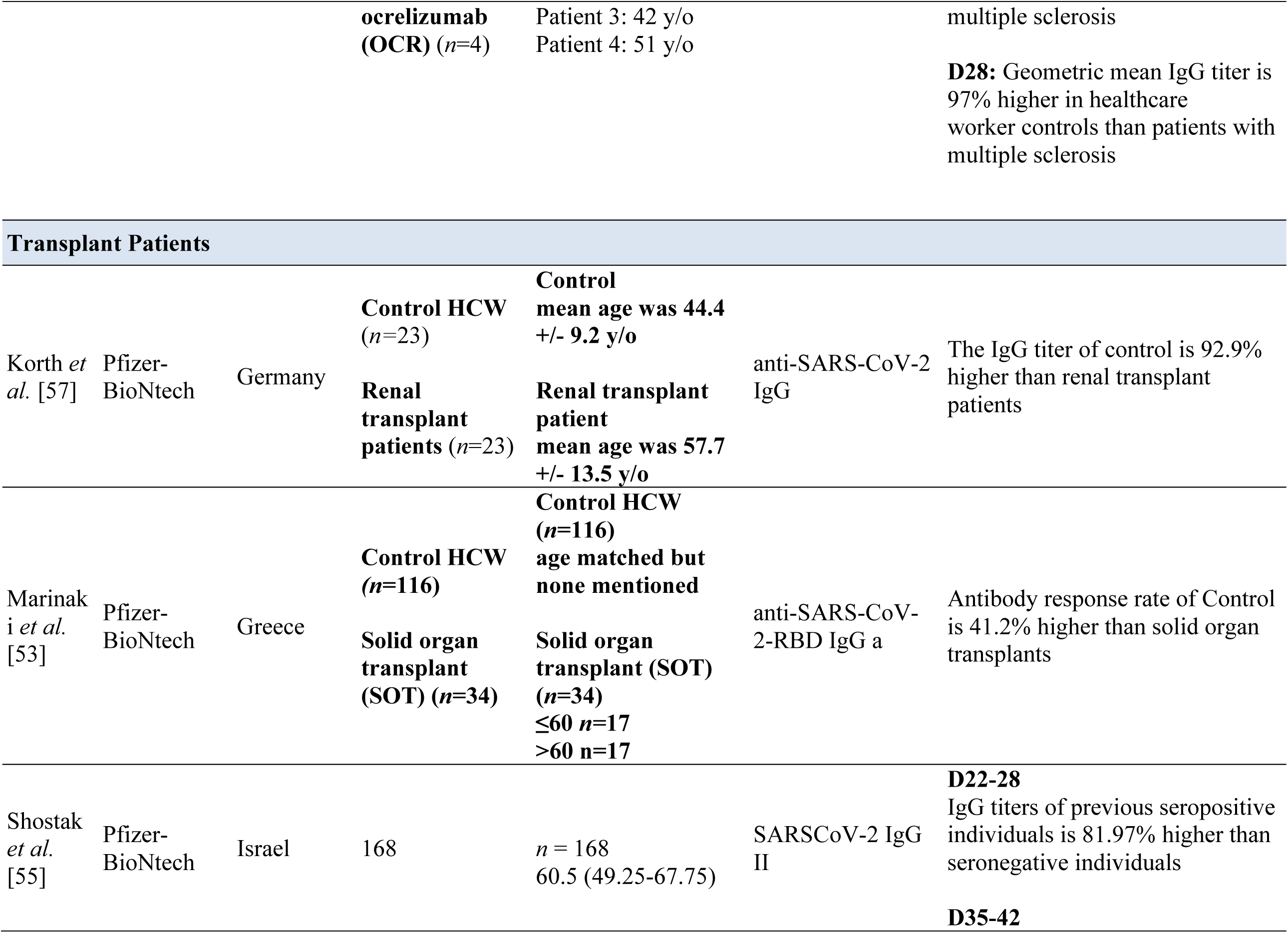
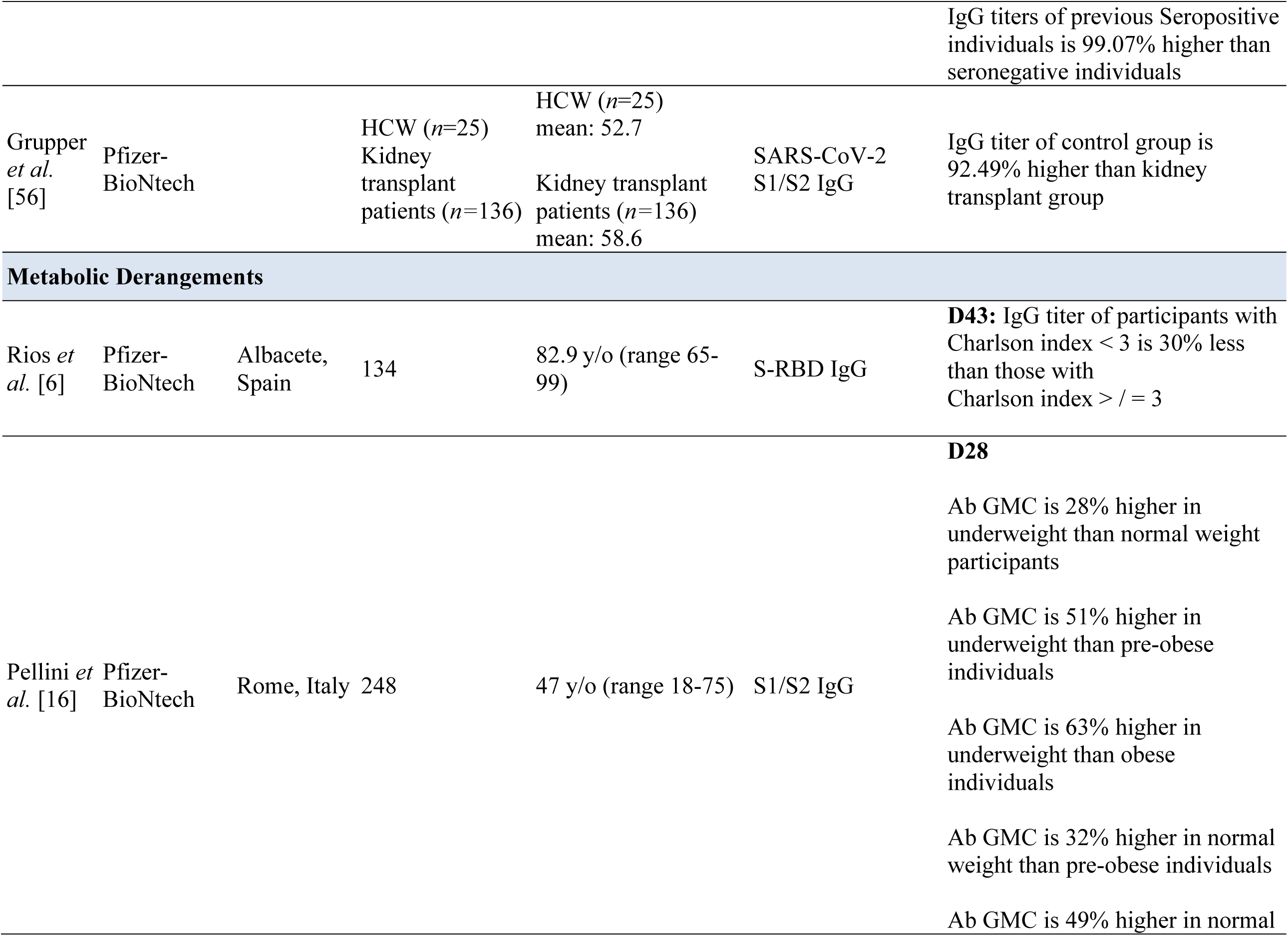
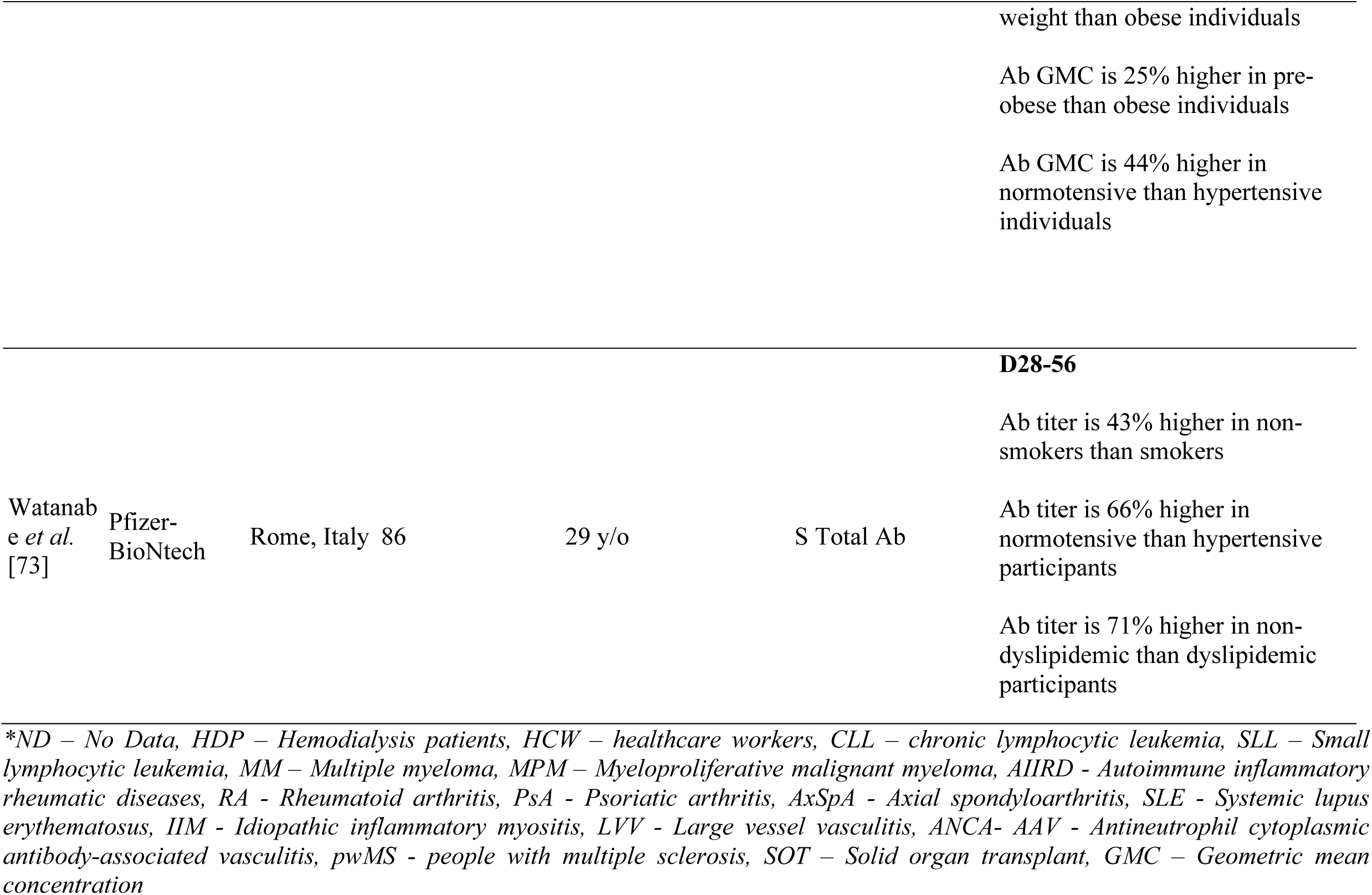
Effect of comorbidities in humoral response following Pfizer-BioNTech (mRNA BNT162b2) vaccine administration.

#### Hemodialysis or End Stage Renal Disease

Hemodialysis patients (HDP) have been identified to be at high-risk of acquiring severe COVID-19 associated symptoms [40]. Most of these patients develop uremia, resulting in ineffective leukocyte function, decreased antigen processing and presentation, and subsequently disrupted innate and adaptive immune responses [41]. In the event of getting infected, these patients cannot fully observe quarantine protocols because of their dependencies on dialysis treatments [42]. Studies have shown that vaccination is the most cost- and resource-effective preventive measure available [43]. We tackled herewith the immunological response of hemodialysis patients to mRNA vaccines.

Among five publications that discussed HDP, only two studies compared HDP to control, and both presented consistent findings that the control group had higher immune responses compared to hemodialysis subjects [43,44]. Nonetheless, all five studies reported a response in majority of their subjects after two doses of the vaccination. Frantzen *et al*. noted that the results go far beyond the hyporesponsive population [45].

Furthermore, Jahn *et al*. reported that HDPs under the age of 60 years responded equally to the control group [43]. However, a limitation of the study was the lack of control subjects over the age of 60 years. In a study by Torregiani *et al*., it was found that younger HDPs (62.31 ± 16.20 years) with lower comorbidity burden are more likely to mount an antibody response and have higher response compared to older patients (73.72 ± 11.18) with more comorbidity burden [44]. This is consistent with the findings of Jahn *et al*. comparing the humoral response of HDPs across different age ranges as presented in Table 4 [43]. Torreggiani *et al*. raised concerns that after the first vaccination only a third of the HDP patients mounted an immune response [46]. Goupil *et al*. also stated that a single dose failed to elicit humoral response among HDP participants with previous SARS-CoV-2 infection and was delayed even in those previously infected [44]. Nevertheless, Goupil *et al*. noted that previously infected patients had higher response compared to the other HDPs.

Overall, there are concerns on lower vaccine efficacy and shorter period of immunoprotection for HDP patients. Suggestions on adjusting the dosage for this group, especially in the elderly population were recommended in all studies.

#### Transplant Recipients

Transplant recipients are at high risk of infection resulting from immunosuppression necessary to prevent organ rejection compounded by immunosuppressive effects of organ failure and chronic disease [47]. Although vaccination has been used as a strategy to prevent infection among transplant recipients, there have been concerns that vaccination may trigger development of donor-specific anti-human leukocyte (HLA) antibodies (DSA) and/or allograft rejection [48,49].

It is thought that vaccination could trigger T- and B-cell responses to vaccine antigens that directly cross-react with alloantigens as in the case of viral infections [50,51]. Furthermore, vaccination can induce cytokine release that may stimulate previously quiescent alloreactive memory responses [50,51]. Adjuvants in vaccines can also lead to non-specific immunostimulating effects that could increase rates of rejection and DSA formation [48,49].

In line with the current pandemic, transplant recipients are a vulnerable group at higher risk for SARS-CoV-2 infection with poorer associated outcomes [52]. The US Centers for Disease Control and Prevention (CDC&P) outlined the safety of the Pfizer-BioNTech and Moderna vaccines because neither vaccine contains live virus that could be dangerous to immunocompromised patients [53]. In this section, we described humoral responses of transplant recipients who have received the mRNA vaccine

Almost all of the studies reviewed had a mean difference of 90% between transplant recipients and control groups. This is because of blunted immune response of transplant patients most likely due to taking immunosuppressive medications. While antibody titers may develop, these usually develop late, are below the protective threshold, and unfortunately wane faster [54,55]. The observed failure to mount appreciable antibody immune response of transplant recipients is consistent in previous findings exploring other common vaccines [56]. Identified contributors to mount a response includes advanced age, need for high dose of corticosteroids during the past 12 months, maintenance with three immunosuppressive medications, and a regimen that includes mycophenolate, antimetabolites or mTOR inhibitors [56].

Korth *et al*. proposed that novel vaccination strategies may be needed to address this failure [57]. This could either indicate more than two booster doses or a combined scheme with mRNA vaccines, protein/subunit vaccines, and vector-based vaccines. Strict evaluation may also be needed after vaccination in the long-term to learn more about the response of this population.

#### Cancer and Autoimmune Disease

Cancer, as a systemic disease, induces functional and compositional changes to the immune system [58]. Some of these changes allow cancer cells to avoid destruction by the immune attack and even support its growth. Studies indicate that B cells responsible for mediating humoral immune response promote and support tumor growth [59]. Depending on the type of cancer, the immunosuppressive tumor microenvironment may vary [60]. In addition, immunosuppression observed among cancer patients may be attributed to their treatment. Similar to the type of cancer, different types of treatment cause different levels of immunosuppression [60]. In a recent study, a significant portion of patients with cancer developed proper humoral immune responses after vaccination [61]. However, recent chemotherapy treatment may be associated with low serologic response [61]. Similar findings were noted on influenza vaccinated patients undergoing chemotherapy compared to healthy controls [62]. Cancer patients who were vaccinated during the early chemotherapy cycle also had better response compared to those in the latter cycle of the treatment [62]. The possible immunosuppression in cancer patients receiving the Pfizer-BioNTech mRNA vaccine was documented by Herishanu *et al*., wherein IgG titer among patients diagnosed with chronic lymphocytic leukemia (CLL) was blunted by 80.65% as compared to controls [63]. Moreover, the same pattern of immunosuppression was reported by Massarweh *et al*. wherein the IgG titer of the control group was 68.48% higher than in cancer patients [64]. Furthermore, similar observations were noted by Pimpinelli *et al*. in myeloproliferative malignancies (MPM) and multiple myeloma (MM) [65]. Response in MPM patients was robust, while MM patients had significantly less response. MM patients undergoing regimens without daratumumab were associated with higher adaptive immune responses. This may be due to daratumumab’s mechanism of action that targets CD38 on the population of normal and tumor plasma cells, thus reducing vaccine immunogenicity by direct depletion of antibody producer cells. Nonetheless, Goshen-lago *et al*. noted that while there was a pronounced lag in antibody production in cancer cases, seroconversion occurred in most patients after the second dose [66].

An autoimmune disease is a condition arising from an abnormal immune system response which mistakenly attacks healthy cells, tissues, and organs. This immune malfunction can affect any part of the body, weakening bodily function that can be potentially fatal [67]. The cornerstone to management of autoimmune disorders is the use of immunosuppressive therapies. However, various immunosuppressive treatments could impact vaccine-induced immunogenicity [68]. For instance, Gallo *et al*. reported that the geometric mean IgG titer of patients with multiple sclerosis (pwMS) treated with ocrelizumab is 97% lower than healthy participants [69]. However, this study is limited by the small number of tested patients and the inability to assess their cell-mediated and innate immune responses. Larger studies exploring the response to SARS-CoV-2 vaccines in pwMS treated with anti-CD20 drugs (*e*.*g*., rituximab) and other high efficacy disease-modifying therapies (DMTs) are necessary to confirm and expand these preliminary data.

Similarly, Furer *et al*. reported an anti-SARS-CoV-2 S1/S2 IgG titer that was 39% lower among participants diagnosed with autoimmune inflammatory rheumatic diseases (AIIRD) as compared to healthy controls [70]. In addition, age could have also been a confounding factor that affected the reported results in this study since majority of the AIIRD participants were elderly with a mean age of 59 years compared to the general population control group with a mean age of 50 years. Among the AIIRDs, rheumatoid arthritis (RA), antineutrophil cytoplasmic antibody (ANCA)-associated vasculitis (AAV), and idiopathic inflammatory myositis (IIM) were associated with a low humoral response to the vaccine which may be partially attributed to their underlying treatment. Furthermore, data presented in this study also have important implications for the management of anti-COVID-19 vaccination in patients with a wide spectrum of AIIRD. Most immunosuppressive treatments, including conventional synthetic disease-modifying antirheumatic drugs (csDMARDs), anticytokine, biologics, and janus kinase inhibitors (JAKi), can be safely continued without significantly attenuating vaccine-induced immunogenicity. In contrast to the recommendation of the American College of Rheumatology, this study does not support withholding methotrexate (MTX) and JAKi in relation to COVID-19 vaccination. Meanwhile, treatment with glucocorticoids, rituximab, abatacept in combination with MTX and mycophenolate mofetil was associated with significantly decreased vaccine-induced immunogenicity. Therefore, timing of vaccination has a critical role in these cases. If clinically feasible, postponing administration of rituximab and abatacept, especially when combined with MTX, seems reasonable to improve vaccine-induced immunogenicity.

#### Metabolic Derangements & Smoking

Metabolic derangement is an important and prevalent comorbidity to consider. Most of which are associated with high body mass index (BMI) and thus, obesity. Recent studies have highlighted the effect of metabolic syndromes on immunity and pathogen defense and coordination of innate and adaptive responses [71]. These changes are associated with decreased immunity from infection, higher risk for complications, and higher rates of vaccine failure [71]. In a study by Sheridan *et al*., BMI values correlated positively with higher initial fold increase in IgG antibodies detected after trivalent influenza vaccine [72]. However, 12 months after vaccination, subjects with higher BMI noted a greater decline in antibody titers. The findings on vaccine failure in relation to BMI and obesity are also noted with mRNA-based vaccines [71]. For instance, Pellini *et al*. reported that there is higher anti-spike S1/S2 IgG production in individuals with lower BMI (BMI <18.5) as compared to pre-obese (BMI 25.0-29.9) or obese (BMI >30.0) participants receiving the Pfizer anti-COVID-19 mRNA vaccine [16]. Furthermore, anti-spike immunoglobulin production was 71% higher in non-dyslipidemic than dyslipidemic participants vaccinated with the Pfizer mRNA [73]. This may be correlated to adipokines, cytokine-like hormones released by adipose tissues that bridge cellular metabolism to immune responses. In particular, leptin plays an important role in controlling the interplay between cellular energy metabolism and regulation of metabolic-immune responses. Indeed, leptin plays a role in modulating cell proliferation, responsiveness, and polarization of T cells. Conversely, leptin promotes B cell homeostasis through inhibition of apoptosis and induction of cell entry. Central leptin resistance is the main risk factor for obesity-related acute and chronic diseases. It also plays a role in dysglycemia, particularly in Type 2 diabetes mellitus as leptin is a therapeutic target for its impact on food intake, body weight and potential to improve insulin action [74].

Hypertension is related to impaired metabolic homeostasis, thus is also regarded as a metabolic disorder [75]. It is important to note that the immune and autonomic systems play an important role in the cause of hypertension and other cardiovascular pathologies [76]. The blunted serologic response noted among hypertensive patients may actually be rooted in a dysfunctional immune system [73]. Both Pellini *et al*. and Watanabe *et al*. reported that IgG production against SARS-CoV-2 in normotensive individuals receiving the mRNA vaccine is significantly higher than in hypertensive participants [16,73].

In addition, ample evidence indicates that cigarette smoking could also affect both innate and adaptive immunity [77]. Cigarette smoking is known to attenuate the normal defensive function of the immune system by affecting the nuclear factor-kappa B (NF-κB) and mitogen-activated protein kinases (MAPKs) signaling, as well as histone modification epigenetics [77]. As reported by Watanabe *et al*., the humoral response of smokers who received the mRNA vaccine was blunted by 43% as compared to non-smokers [73]. Based on these studies, it is increasingly clear that suboptimal metabolic health and unhealthy lifestyle practices are associated with poor vaccine-induced immunogenicity.

## Conclusion

Humoral immune response differs in every individual and is affected by many factors such as age, sex, serostatus, and underlying comorbidities. This systematic review showed that old individuals (>65 years) produce lower anti-SARS-CoV-2 antibody levels and had higher chances to be low- or non-responders, especially when combined with other comorbidities. Interestingly, the female sex has greater antibody production due to the immunomodulating properties of estrogen and the X chromosome. Moreover, due to early antibody response present among seropositive individuals, higher levels of antibodies were measured post-vaccination than seronegative individuals. Presence of comorbidities also showed significant decline in antibody production especially if present in the elderly population. Hemodialysis, transplantation, cancer and autoimmune diseases, as well as metabolic derangements, all shared a blunted humoral immune response which could be rooted to dysfunctional immune system and several factors such as aging, which serves as a significant aggravating factor, as well as the use of immunosuppressive and antimetabolite medications. Hence, further studies in improving vaccination strategies for COVID-19 should be employed in consideration of the aforementioned factors.

It is worth mentioning that additional mRNA-based vaccine boosters have already been attempted in some of these populations of low responders. While presenting a relatively safe reactogenic response comparable to that seen after the first or second mRNA vaccine dose [79], the third vaccine dose was found to be effective in further boosting antibody levels in elderly people [80], in solid organ transplant recipients [81], and in patients receiving maintenance hemodialysis or peritoneal dialysis [82]. The third dose was also effective in reinforcing immunity against VoC [83]. Irrespective of the demographical or clinical conditions that would blunt the anti-SARS-CoV-2 antibody response, the importance of monitoring humoral immunity seems almost unquestionable for prioritizing vaccine boosters, including new vaccines able to efficiently protect against current and potentially future SARS-CoV-2 drifted VoC. Their administration could provide the best compromise between the still limited availability and the highest clinical efficacy in averting or limiting COVID-19 infections and/or severe illness in especially vulnerable populations [84].

## Data Availability

All data produced in the present work are contained in the manuscript.

## Recommendations

For future researchers it is recommended to investigate further on the correlation and interdependence after combining age, sex, serostatus, and comorbidities and their effect on humoral response. In addition, comparison between other SARS-CoV-2 mRNA vaccines such as Moderna (mRNA 1273) with Pfizer-BioNTech (mRNA BNT162b2) could also be explored using these demographic parameters.

